# Estimating the extent of asymptomatic COVID-19 and its potential for community transmission: systematic review and meta-analysis

**DOI:** 10.1101/2020.05.10.20097543

**Authors:** Oyungerel Byambasuren, Magnolia Cardona, Katy Bell, Justin Clark, Mary-Louise McLaws, Paul Glasziou

## Abstract

**Background:** The prevalence of true asymptomatic COVID-19 cases is critical to policy makers considering the effectiveness of mitigation measures against the SARS-CoV-2 pandemic. We aimed to synthesize all available research on the asymptomatic rates and transmission rates where possible.

**Methods:** We searched PubMed, Embase, Cochrane COVID-19 trials, and Europe PMC (which covers pre-print platforms such as MedRxiv). We included primary studies reporting on asymptomatic prevalence where: (a) the sample frame includes at-risk population, and (b) there was sufficiently long follow up to identify pre-symptomatic cases. Meta-analysis used fixed effect and random effects models. We assessed risk of bias by combination of questions adapted from risk of bias tools for prevalence and diagnostic accuracy studies.

**Results:** We screened 2,454 articles and included 13 low risk-of-bias studies from seven countries that tested 21,708 at-risk people, of which 663 were positive and 111 were asymptomatic. Diagnosis in all studies was confirmed using a RT-PCR test. The proportion of asymptomatic cases ranged from 4% to 41%. Meta-analysis (fixed effect) found that the proportion of asymptomatic cases was 17% (95% CI: 14% - 20%) overall; higher in aged care 20% (14% - 27%), and lower in non-aged care 16% (13% - 20%). Five studies provided direct evidence of forward transmission of the infection by asymptomatic cases. Overall, there was a 42% lower relative risk of asymptomatic transmission compared to symptomatic transmission (combined Relative Risk: 0.58; 95% CI 0.335-0.994, p=0.047).

**Discussion:** Our estimates of the prevalence of asymptomatic COVID-19 cases and asymptomatic transmission rates are lower than many highly publicized studies, but still sufficient to warrant policy attention. Further robust epidemiological evidence is urgently needed, including in sub-populations such as children, to better understand the importance of asymptomatic cases for driving spread of the pandemic.

**Funding:** OB is supported by NHMRC Grant APP1106452. PG is supported by NHMRC Australian Fellowship grant 1080042. KB is supported by NHMRC Investigator grant 1174523. All authors had full access to all data and agreed to final manuscript to be submitted for publication. There was no funding source for this study.

## Introduction

Asymptomatic cases of any infection are of considerable concern for public health policies to manage epidemics. Such asymptomatic cases complicate the tracking of the epidemic, and prevent reliable estimates of transmission, tracing, and tracking strategies for containing an epidemic by isolating and quarantining. This has been a significant concern for the current COVID-19 pandemic.(1)

The possibility of asymptomatic transmission of COVID-19 cases was first raised by a case report in China where a traveler from Wuhan was presumed to have transmitted the infection to 5 other family members in other locations while she remained asymptomatic for the entire 21-day follow-up period.(2) Subsequently a number of other reports confirmed not only the possibility but began quantifying the potential proportions. For example, the outbreak on the Diamond Princess cruise ship(3) demonstrated a substantial proportion of asymptomatic cases after widespread testing of those on board the ship. An early rapid review by the Centre for Evidence Based medicine in Oxford(4) found that the estimated proportion of asymptomatic COVID-19 cases ranged from 5% to 80%. However, many of the identified studies were either poorly executed or poorly documented, making the validity of these estimates questionable.

We therefore sought to identify all studies that had attempted to estimate the proportion of asymptomatic COVID-19 cases, select those with low risk of bias, and synthesize these to provide an overall estimate and potential range. We also aimed to estimate the rate of forward transmission from asymptomatic cases if sufficient data were found.

## Methods

We conducted a systematic review and a meta-analysis using enhanced processes with initial report completed within two weeks, using daily short team meetings to review the progress, plan next actions, and solve discrepancies and other obstacles.(5) We also used locally developed open access automation tools and programs such as the Polyglot Search Translator, SearchRefiner, and the SRA Helper to design, refine and convert our search strategy for all the databases we searched and to speed up the screening process.(6) We searched PROSPERO database to rule out existence of a similar review; searched PubMed, Embase, Cochrane COVID-19 trials for published studies, and Europe PMC for pre-prints from January 2019 to 20 Jul 2020. A search string composed of MeSH terms and words was developed in PubMed, and was translated to be run in other databases using the Polyglot Search Translator. The search strategies for all databases are presented in Supplement 1. We also conducted forward and backward citation searches of the included studies in the Scopus citation database.

We restricted publication types to reports of primary data collection released in full (including pre-prints) with sufficient details to enable a risk of bias assessment and contacted authors for clarifications on follow-up times and sampling frames. We anticipated cross-sectional prevalence surveys with follow up, and cohort studies would be the bulk of eligible reports. No restrictions on language were imposed. We excluded studies for following reasons: sampling frame in part determined by presence or absence of symptoms; no or unclear follow up; no data on asymptomatic cases; single case study/small cluster; modelling or simulation studies (but sources of real data were checked for possible inclusion); non-SARS-CoV-2 virus study; antiviral treatment studies; study protocols, guidelines, editorials or historical accounts without data to calculate primary outcomes.

### Participants

We included studies of people of any age where: all those at-risk of contracting SARS-CoV-2 virus were tested regardless of presence or absence of symptoms; diagnosis was confirmed by a positive result on a real-time reverse transcription polymerase chain reaction (RT-qPCR); and all cases had a follow up period of at least 7 days to distinguish asymptomatic cases from pre-symptomatic cases (Figure 1).

**Figure 1.**
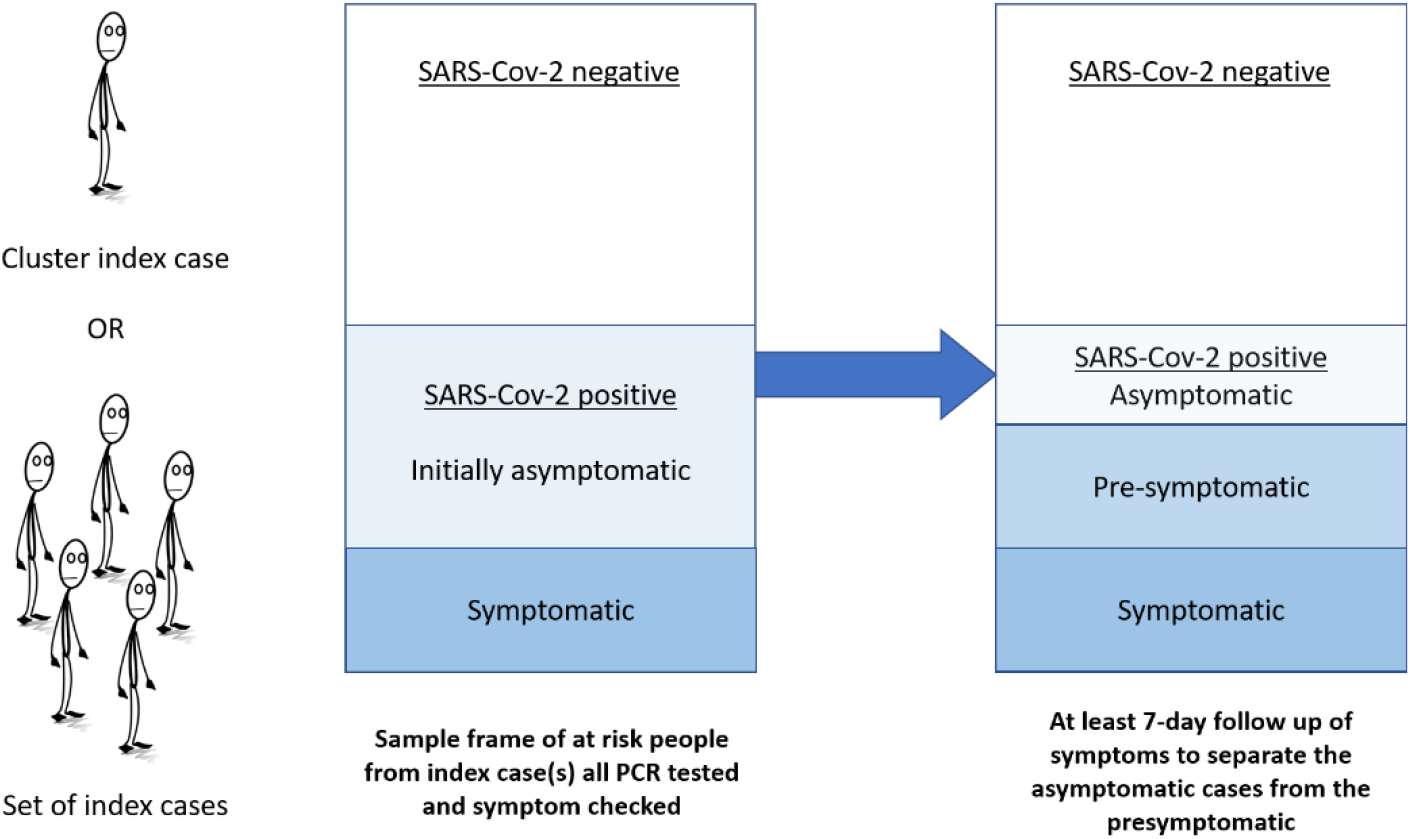
Depiction of ideal study flow and criteria used for study inclusion. (i) sample frame of at-risk people, and (ii) adequate follow-up on symptoms.

### Outcomes

Our primary outcome was proportion of all people with SARS-Cov-2 infection who were completely asymptomatic at the time of test and throughout the follow up period, where the denominator included all tested individuals in the study sample whose result was positive, and the numerator included those who tested positive and had no symptoms. Our secondary outcome was estimate of onward transmission of the infection from asymptomatic cases.

### Study selection and screening

Two authors (OB and MC) independently screened titles, abstracts, and full texts according to eligibility criteria. All discrepancies were resolved via group discussion with the other authors. Reasons for exclusion were documented for all full text articles deemed ineligible (Supplement 2) - see PRISMA diagram (Figure 2).

### Data extraction

Three authors (OB, MC, KB) used a Microsoft Excel form to extract the following information:

1. Methods: study authors, year of publication, country, publication type, duration of study, duration of follow-up
2. Participants: sample size, age (mean or median; range), setting (community, province, aged care facility, hospital, screening clinic), presence or absence of symptoms, test results.
3. History of illness and diagnosis: Type of test, numerator, denominator/sampling frame, proportion of asymptomatic, mild symptomatic, or symptomatic subjects, and number or proportion of people infected by the asymptomatic case.

Case definition: Asymptomatic: confirmed via any testing specified above with report of no symptoms for the duration of sufficient follow-up to differentiate from pre-symptomatic cases. Exposure: contact with a confirmed case or potential contact of another pre-symptomatic person (e.g. came from an endemic area or linked with an infected traveler). The World Health Organization (WHO) recommends that “for confirmed asymptomatic cases, the period of contact is measured as the 2 days before through the 14 days after the date on which the sample was taken which led to confirmation”.(7)

### Risk of bias assessment

Three authors (OB, MC, KB) assessed the risk of bias of potentially includable studies. We used a combination of risk of bias tools for prevalence studies(8) and diagnostic accuracy(9) and adapted the key signaling questions on sampling frame, ascertainment of infectious disease status, acceptability of methods to identify denominators, case definition of asymptomatic for the numerator, and length of follow up, as shown in Table 2 and in Supplement 3 in full.

### Data analysis

We estimated the proportion of COVID-19 cases who were asymptomatic for each included study population, assuming a binomial distribution and calculating exact Clopper–Pearson confidence intervals. We then pooled data from all included studies using (1): fixed effects meta-analysis and (2): random effects meta-analysis. All analyses were conducted using SAS 9.4; the FREQ procedure was used for individual studies and the fixed effect meta-analysis; the NLMIXED procedure was used for the random effects meta-analysis.

We also meta-analyzed the forward transmission rates from asymptomatic and symptomatic cases where there was sufficient data and report the pooled relative risk comparing the two. We planned to undertake subgroup analysis for age (between studies, and within studies where age was reported separately for asymptomatic and symptomatic cases). As only studies deemed to be of high quality on items 1 and 2 after risk of bias appraisal were included in the analysis, no sensitivity analysis of high versus low quality studies was undertaken. Instead, we did a sensitivity analysis where we omitted studies with follow-up duration of less than 14 days.

## Results

Two thousand four hundred and fifty-four articles were screened for title and abstract and 161 full-text articles assessed for inclusion (Figure 2). Major reasons for exclusion were inadequate sampling frame and insufficient follow-up time to accurately classify the asymptomatic cases. Full list of excluded studies with reasons is presented in Supplement 2. Thirteen articles − nine published and four preprints − from seven countries (China (4), United States of America (USA) (4), Taiwan (1), Brunei (1), Korea (1), France (1) and Italy (1)) that tested 21,708 close contacts of at least 849 confirmed COVID-19 cases, of which 663 were positive and 111 were asymptomatic, met eligibility criteria for the estimation of the primary outcome. (10-22)

**Figure 2.**
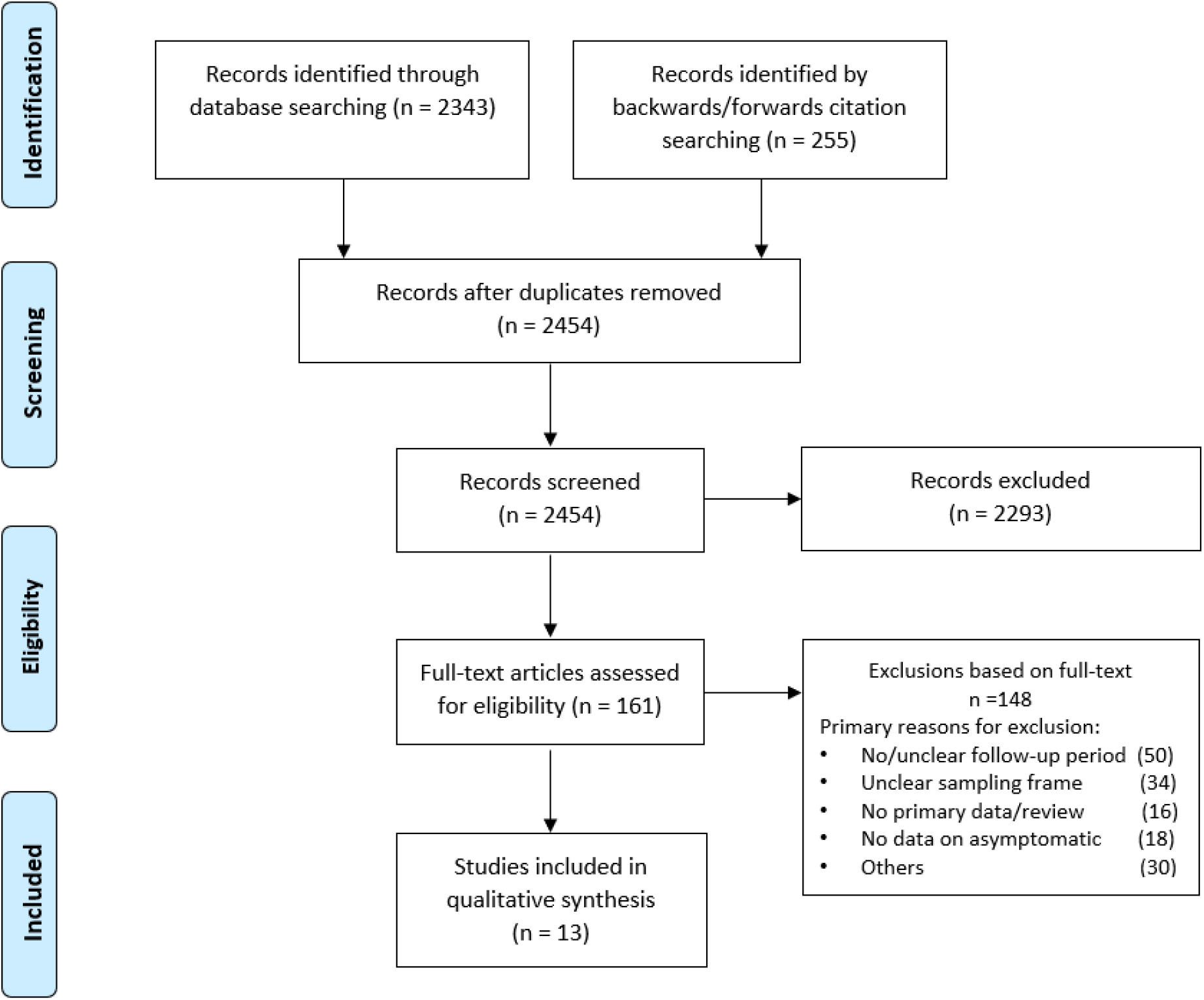
Screening and selection of articles.

Their sampling frames included residents of skilled nursing facilities (SNF) (10, 12, 15, 19, 20); high-risk close contacts of confirmed COVID-19 cases (11, 13, 14, 17, 18, 21); and a whole district surveillance program in Italy (16). The demographic characteristics (Table 1) indicate that most of the tested individuals were adults, with mean age over 75 years in the five SNF studies and mean over 31 years in the non-aged care studies. The proportions of children and young people (0-20 years) ranged from 6% to 23.5%.

Diagnosis in all studies was confirmed via RT-PCR and in two cases supplemented with radiological evidence(17, 21). Testing of individuals within the study sample varied across settings but was generally very high: all contacts regardless of symptoms(11, 13, 14, 17, 18, 21); >97% of SNF residents(10, 12, 15, 19, 20) and 85.9% of an entire town(16). Length of follow-up for monitored individuals in the SNF studies was 7-30 days(10, 12, 15, 19, 20); 14 days for the Bruneian(13), Taiwanese(14), Korean(18) and Chinese close contacts(17, 22); 7-14 days in the Italian community(16); 12 days for 95% of all contacts in the Shenzhen community surveillance(11), and 16±6 days in Liaocheng, China(21).

**Table 1.** Characteristics of included studies (n=13)

| **Study ID, country, and publication status** | **Study population (sampling frame)** | **Sample size, age** | **Diagnostic testing and frequency** | **Length of follow up of asymptomatic cases** |
| --- | --- | --- | --- | --- |
| **Roxby et al (USA)**<br>Published | Residents of independent and assisted living communities (Facility 1) in Seattle following two confirmed cases between 5-9 Mar. | N=79. Mean age of cohort 86 years. | Nasal swab, RT-PCR, twice a week apart | 7 days |
| **Patel et al (USA)**<br>Published | Residents and staff of skilled nursing facility in Illinois on 15 Mar. | n=126. Median age of cases 82 years. | Nasal swab, RT-PCR, once | 30 days |
| **Dora et al (USA)**<br>Published | Residents of skilled nursing facility in Veterans Affairs Greater Los Angeles Healthcare System between 29 Mar-23 Apr. | N=99. Median age of cohort 75 years. | Nasal swab, RT-PCR, repeated every 10 ds | At least 14 days |
| **Blain et al (France)**<br>Published | Nursing home residents in France tested weekly since early Mar. | N=79. Mean age 86 years. | Nasal swab, RT-PCR, repeated weekly | 6 weeks |
| **Arons et al (USA)**<br>Published | Residents of skilled nursing facility (Facility A) in Seattle following a confirmed case on 1 Mar. | N=86. Mean age of cohort 77 years, mean age of cases 79 years. | Nasal swab, RT-PCR, twice a week apart | 7 days |
| **Zhang et al (China)**<br>Published | Close contacts of confirmed cases between 28 Jan -15 Mar in Guangzhou, China. | N=369. Median age 35 years. | Nasal swab, RT-PCR, at least twice | 14 days |
| **Tian et al (China)**<br>Preprint | Close contacts (co-workers, family members, customers) of a confirmed supermarket employee (super-spreader) in Liaocheng city, China. | N~8000. Mean age of cases 48 years. | Nasal swab, RT-PCR, repeated every 2 days | 16±6.15 days |
| **Cheng et al (Taiwan)**<br>Published | High risk close contacts (household members, HCWs) of first 100 cases in Taiwan. | N=849. Mean age of cohort 42 years, mean age of cases 41 years. | Nasal swab, RT-PCR, repeated during 14-d quarantine | 14 days |
| **Lavezzo et al (Italy)**<br>Published | Majority of population of Italian town of Vo following a COVID-19 death on 21 Feb. | N=2,812. Mean age of cohort 47 years, mean age of cases 58 years. | Nasal swab, RT-PCR, twice, 7-14 ds apart | 7-14 days |
| **Bi et al (China)**<br>Published | Close contacts of cases confirmed before 9 Feb in Shenzhen, China. | N=1,286. Mean age of cohort 38 years, mean age of cases 43 years. | Nasal swab, RT-PCR, repeated during 14-d quarantine | 95% followed up for 12+ days |
| **Chaw et al (Brunei)**<br>Preprint | Bruneian attendants of a religious event in Malaysia, where a confirmed case was present. | N=1830. Mean age of cohort 31 years, mean age of cases 33 years. | Nasal swab, RT-PCR, repeated weekly | 14 days |
| **Luo et al (China)**<br>Preprint | Close contacts of 347 confirmed COVID-19 patients identified between 13 Jan and 6 Mar in Guangzhou, China. | N=4,950. Mean age of cohort 38 years, mean age of cases 44 years. | Nasal swab, RT-PCR, repeated every 2 ds | 14 days |
| **Park et al (Korea)**<br>Published | Employees, residents, and visitors of a commercial+ residential building where a confirmed case worked. | N=1143. Mean age of cohort 38 years. | Nasal swab, RT-PCR, repeated during 14-d quarantine | 14 days |

RT-PCR: Reverse Transcription-Polymerase Chain Reaction; HCWs: health care workers.

The proportion of asymptomatic cases in the 13 included studies ranged from 4% (95% CI 1% - 10%) in Korea(18) to 40% in Vo, Italy (16) and in an aged care facility in USA(20). Combining data from all 13 studies, we estimate that 17% of cases were asymptomatic (95% CI: 14% - 20%; fixed effects); for the eight non-aged care studies: 16% (13% - 19%), and for the five studies of SNFs 20% (14% - 27%) (Figure 3). The corresponding estimated proportions in the random effects meta-analysis were: overall 18% (95% CI: 9% - 26%), non-aged care 16% (7% - 26%), and aged care 21% (5% - 36%). The 95% prediction interval was 4%-52%. In the sensitivity analysis omitting studies where length of follow up was less than 14 days (10, 11, 16, 20), the overall estimate was modestly lower at 15% (fixed effect, 95% CI: 12%-18%) or 17% (random effects, 95% CI: 8-26%). Heterogeneity as expressed by I^2^ was 84%.

**Figure 3.**
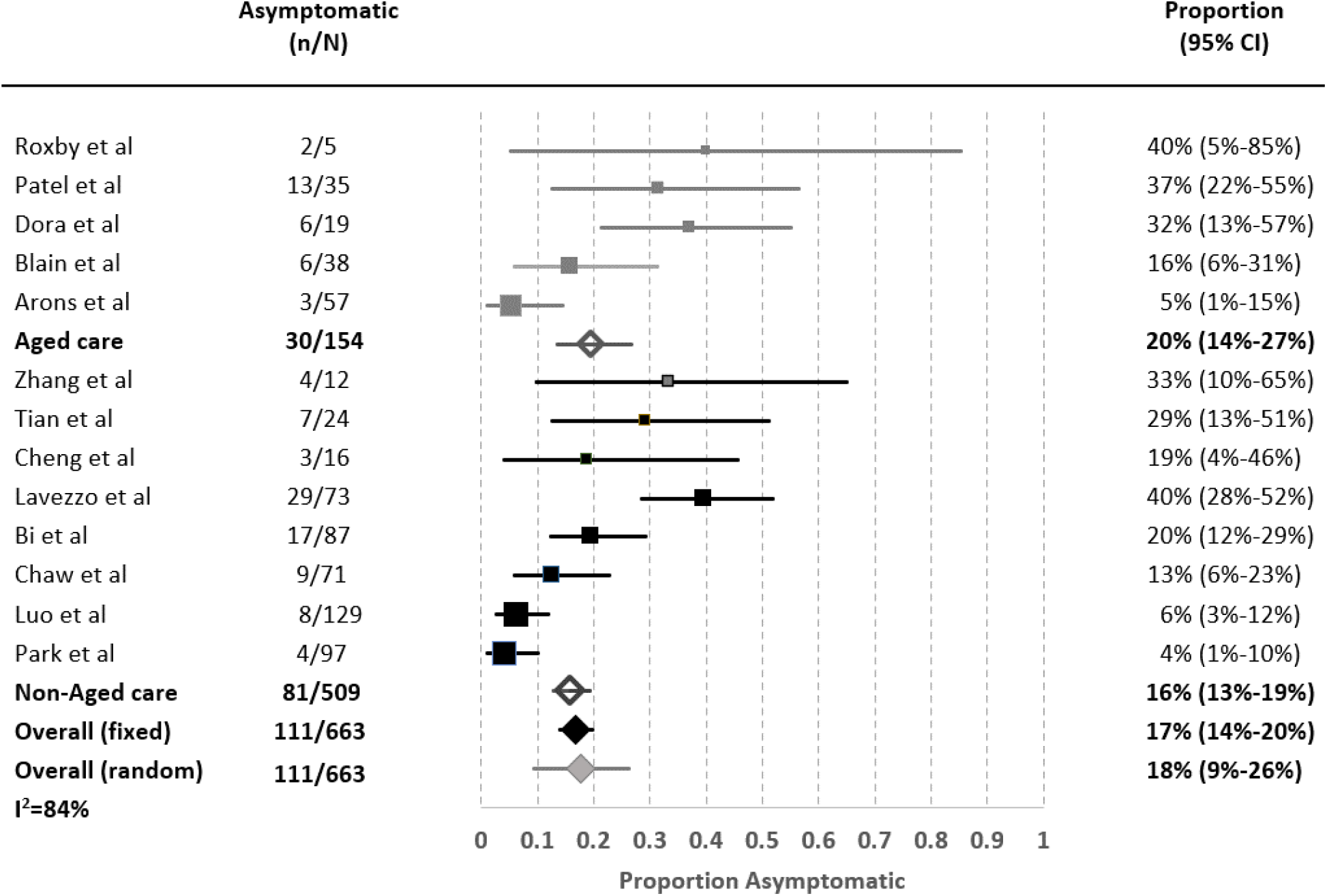
Pooled estimates of proportion of asymptomatic carriers by subpopulations. N - positive cases; n - asymptomatic cases.

Five studies reported data on secondary infection transmission from asymptomatic cases (Table 2). The asymptomatic transmission rates ranged from none to 2.2%, whereas symptomatic cases’ transmission rates ranged between 0.8-15.4%. Cycle threshold (Ct) from real-time RT-PCR assays or the viral load did not differ between asymptomatic and symptomatic individuals in three of the studies.(10, 14, 16) Overall there was a 42% lower relative risk of asymptomatic transmission compared to symptomatic transmission (pooled Relative Risk: 0.58; (fixed-effects 95% CI 0.335-0.994, p=0.047); 0.38 (random-effects 95% CI 0.13-1.083, p=0.07). I^2^=43.4%).

**Table 2.**
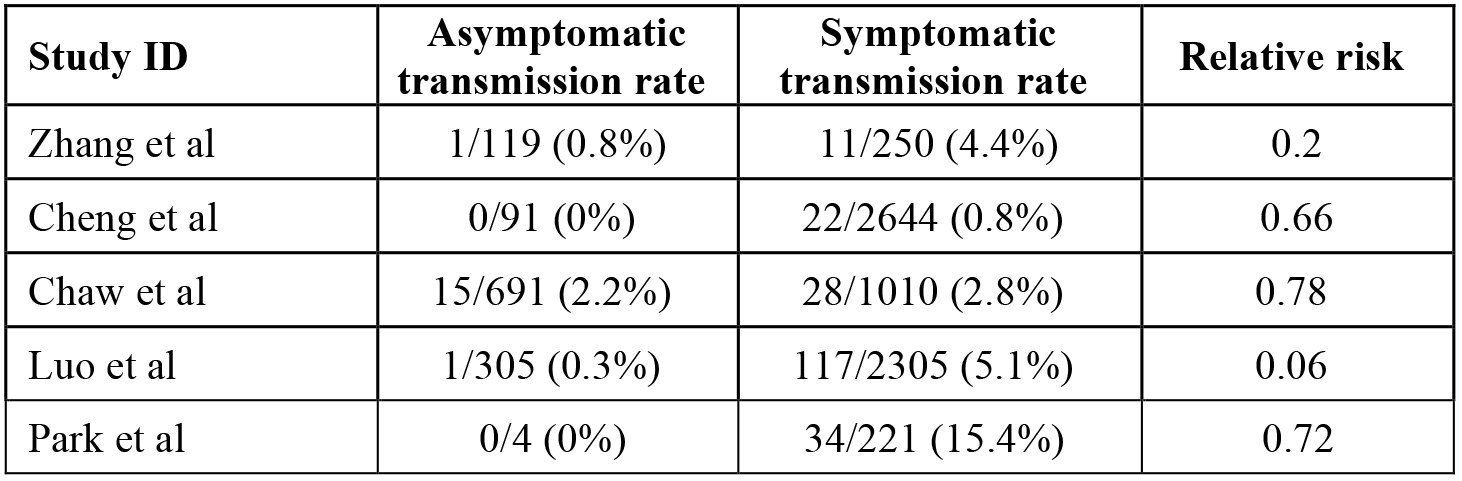
Comparison of secondary transmission rates.

### Risk of bias of included studies

Table 3 summarizes the overall risk of bias assessment of the nine included studies (full list of risk of bias questions in Supplement 3). All of the studies were evaluated as low risk of bias for the sampling frame and length of follow up domains, which were part of the inclusion criteria (Domain 1 and 5). Two studies had potential non-response bias for not testing all of the eligible participants (14% (463/3275) of the target population was not tested in Lavezzo et al study(16)) or not reporting all tested participants results (87/98 cases were reported in Bi et al study(11)) (Domain 2). Four studies either had not tested the study population at least twice during the follow up period or had not provided clear information on it (Domain 3). Nine studies did not explicitly state the asymptomatic case definition they adhered to or had additional bias due to high percentage of people in the SNFs with severe cognitive impairment(10, 12, 15, 19, 20)(Domain 4).

**Table 3.**
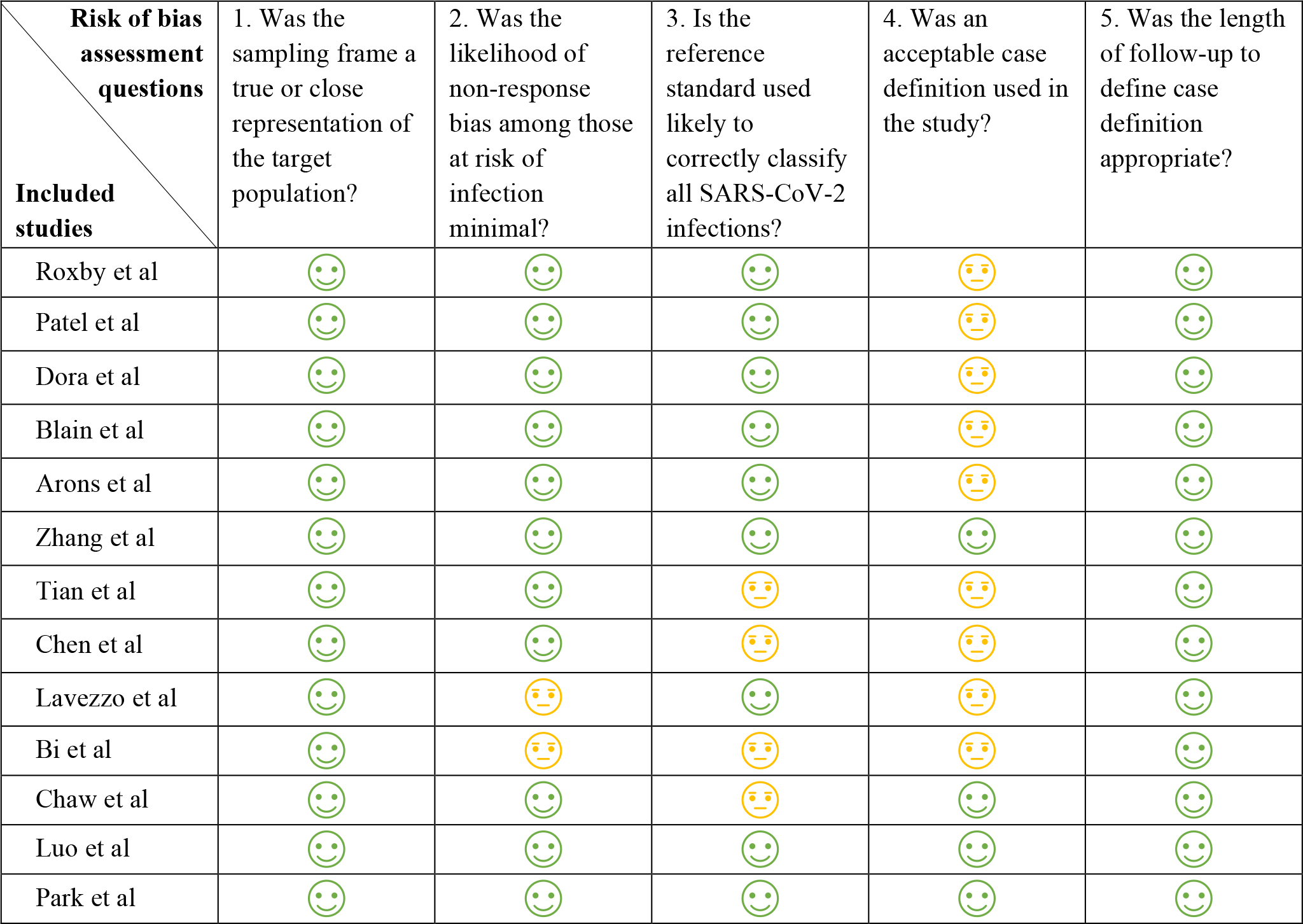
Risk of bias in 9 included studies. Green smiley face denotes low risk, yellow straight face – moderate or unclear risk.

### Excluded studies

Several well publicized studies did not meet our inclusion criteria. The outbreak on the Diamond Princess cruise ship involved 3,711 passengers of whom over 600 acquired COVID-19.(3) Many of the positive cases were relocated to medical facilities in Japan without details of their clinical progression. To correct for the lack of follow-up, Mizumoto and colleagues applied a statistical adjustment for the right censoring and estimated that 17.9% (95% CI 15.5% - 20.2%) of positive cases were asymptomatic.

An open invitation screening of the Icelandic population suggested around 0.8% of the population were SARS-CoV-2 positive, with half classified as (initially) asymptomatic.(2) However, as there was no follow-up, we cannot separate asymptomatic from pre-symptomatic. Furthermore, the study excluded symptomatic people undergoing targeted testing, which impeded an estimate of an overall asymptomatic rate.

A study of 215 pregnant women in New York identified 33 SARS-CoV-2 positive women.(23) On admission to the delivery unit, 4 of the 33 positive cases were symptomatic and 3 became symptomatic before postpartum discharge, suggesting an asymptomatic rate of 26/33 (79%). However, the 2 days of follow-up was insufficient to meet our inclusion criteria.

A case report of a pre-symptomatic Chinese businessman transmitting COVID-19 to a German business partner was also excluded because despite three other people acquiring the infection from the affected German source, none of them was asymptomatic at follow-up.(24) A 5-day point-prevalence testing of adults living in homeless shelters in Boston found 147 positive cases of which “the majority” had mild or no symptoms.(25) We excluded this study, as there was no numeric estimate for true asymptomatic, and no follow-up assessment.

Two studies examined people repatriated from overseas to their home countries by plane. Neither study was clear whether symptomatic people could board the plane and be included - and if excluded, they would overestimate the asymptomatic rates. The study of 565 Japanese citizens repatriated from China,(26) found 13 positives: 4 asymptomatic and 9 symptomatic, based on screening on arrival. The other of 383 Greek citizens repatriated from UK, Spain, and Turkey(27) found 40 asymptomatic positives on arrival, 4 of whom later self-reported symptoms. Again, the likely initial exclusion of symptomatic people, and the lack of comprehensive follow up would both overestimate the asymptomatic rates.

## Discussion

Though the rate of asymptomatic COVID-19 cases has received considerable attention, we could find only 13 studies that provided an adequate sample frame and follow-up to ascertain a valid estimate of the proportion of asymptomatic cases. The combined estimate of the asymptomatic proportion was 17% (95% CI 14% - 20%), but with considerable heterogeneity (I^2^=84%) and a 95% prediction interval that ranged from 4% to 52%. There was no clear difference in the proportions between aged care and non-aged care studies. Only five of the 13 studies provided data on transmission rates from asymptomatic cases. The transmission risk from asymptomatic cases appeared to be lower than that of symptomatic cases, but there was considerable uncertainty in the extent of this (RR 0.58; 95% CI 0.335-0.994, p=0.047).

Strengths of our systematic review include achieving full methodological rigor within a much shorter time frame than traditional reviews using enhanced processes and automation tools.(5) We also critically assessed the risk of bias of all full text articles we screened to include studies with the least risk of bias in sampling frame and length of follow up domains to be able to differentiate between the asymptomatic and pre-symptomatic cases.

There are several limitations to our findings. First, our search focused on published and pre-print articles, and may have missed some public health reports that are either unpublished or only available on organisational websites. Second, the design and reporting of most of the studies had a number of important deficits that could impact their inclusion or our estimates. These deficits include the poor reporting of the sample frame, the testing and symptom check, and the follow-up processes. Such reporting would have been considerably aided by a flow chart of cases (as Lavezzo et al does) of identification, testing, and follow-up including missing data. A further important limitation was the poor reporting of symptoms, which was often simply dichotomised into symptomatic versus asymptomatic without clear definitions and details of possible mild symptoms. The included studies did not report sufficient data to examine the impact of age and underlying comorbidities on the asymptomatic rate. Finally, all included studies relied on RT-qPCR, hence some cases might have been missed due to false negative result, especially where study participants were only tested once.(28) If the tests missed more asymptomatic cases, then the true proportion of asymptomatic cases could be higher than our estimates. On the other hand, false positive results which may occur when people without symptoms are tested in low prevalence settings, would mean the true prevalence of asymptomatic cases was lower than our estimates.

Several other non-systematic and systematic reviews have examined the proportion of asymptomatic cases. The non-systematic reviews estimated asymptomatic rates between 5% and 80%.(4, 29) However, they only included early cross-sectional reports and did not critically appraise the study design, nor attempted to pool the most valid studies. Five other systematic reviews reported pooled estimates of asymptomatic rate between 8%-16%.(30-34) However, these reviews included studies we excluded due to high risk of bias in sampling frame. Ongoing monitoring for new studies is warranted but should include robust methodological assessment including ensuring included studies have sufficient follow up period to differentiate the asymptomatic from presymptomatic cases. Our review currently also has a more recent search date than the other reviews and includes sensitivity analysis by length of follow up time. Our estimate on risk of transmission by asymptomatic cases was comparable to two other empirical data reported by Buitrago-Garcia et al (RR 0.35) and Koh et al (RR 0.39).(32, 34)

Estimates of the proportion of the cases that are asymptomatic and the risk of transmission are vital parameters for modelling studies. Our estimates of the proportion of asymptomatic cases and their risk of transmission suggest that asymptomatic spread is unlikely to be a major driver of clusters or community transmission of infection, but the extent of transmission risk for pre-symptomatic and minor symptomatic cases remains unknown. The generalisability of the overall estimate is unclear, and we observed considerable variation across the included studies which had different settings, countries, and study design, reflected in the reasonably wide prediction interval.

Many unanswered questions about asymptomatic cases remain. Only one of the more recent studies we included tested the patients for IgG antibodies to determine the seroconversion in the elderly. Without repeated and widespread RT-PCR and antibody tests, it is difficult to accurately estimate the prevalence of COVID-19 infection and inform our infection prevention strategies for.(35) The role of viral load and virus shedding dynamics in asymptomatic and symptomatic cases will further help answer the question of forward transmission and disease length and severity. Other unknowns include whether there is a difference in in the proportion of cases that are asymptomatic according to age (particularly children vs adults), sex, or underlying comorbidities; and whether asymptomatic cases develop long-term immunity to new infections. For most studies, the PCR (+) cases were traced from the index cases and the testing were carried out mostly at the beginning of the pandemic wave for the locale. So, for this review of inception cohorts, people with long-term persistent positive testing were unlikely to be misclassified as asymptomatic. The issue of persistent PCR positivity after a person has recovered from infection might be of concern to more recent studies conducted at some time after the “first” wave of the pandemic has happened. In such studies, researchers will need to ask about history of illness compatible with COVID-19 even if this occurred months ago, and the PCR testing could be supplemented by other tests such as viral culture and anti-SARS-CoV-2 antibody tests.

Our recommendations for future research also include improved clearer reporting of methods, sampling frames, case definition of asymptomatic, extent of contact tracing, duration of follow-up periods, presentation of age distribution of asymptomatic cases and separation of presymptomatic and mild cases from asymptomatic cases in result tables. Most studies used a limited definition of asymptomatic COVID-19 case. That could lead to mixing paucisymptomatic people with the asymptomatic cases. If that were common issue, then the true asymptomatic prevalence would be even lower than the current estimates. A reliable estimate of the proportion of asymptomatic cases and the burden of disease is imperative in our understanding of infection transmission capacity of asymptomatic cases to inform public health measures for these individuals who according to our findings appear to pose lower risk of transmission. Until we have further immunological and epidemiological evidence, we advise that the importance of asymptomatic cases for driving the spread of pandemic to be considered with caution.

## Data Availability

Further data can be requested from the authors.

## Authors’ contributions

PG conceived the study and co-designed with OB, MC, and KB. JC led the literature searches including backward and forward citation analysis. OB and MC conducted the parallel title, abstract and full text screening. OB, MC, PG, KB did data extraction. KB and PG did meta-analysis. OB, MC and KB did risk of bias analysis of included studies. MLM provided expertise in interpretation of the findings. All authors contributed to resolving disagreements throughout the study conduct and to writing of the manuscript.

## Conflict of interest

All authors have completed the ICMJE uniform disclosure form. Prof Mary-Louise McLaws is a member of World Health Organization (WHO) Health Emergencies Program Experts Advisory Panel for Infection Prevention and Control (IPC) Preparedness, Readiness and Response to COVID-19 and WHO IPC Guidance Development Group for COVID-19.

## Acknowledgment

We thank the authors of eligible manuscripts for their replies to our queries.

## Notes

### Funding Statement

No external funding was received for this work.

### Author Declarations

Ethics approval was not required.

### Summary of Updates

This version has the most uptodate search date of 20 Jul 2020. We added four new studies to the meta-analysis and the current 13 high-quality study estimate of asymptomatic COVID-19 is 17% (95% CI: 14%-20%). There was also 42% lower relative risk of asymptomatic transmission compared to symptomatic transmission (combined Relative Risk: 0.58; 95% CI 0.335-0.994, p=0.047).

